# Safety of ‘hot’ and ‘cold’ site admissions within a high volume urology department in the United Kingdom at the peak of the COVID-19 pandemic

**DOI:** 10.1101/2020.08.04.20154203

**Authors:** Luke Stroman, Beth Russell, Pinky Kotecha, Anastasia Kantartzi, Luis Ribeiro, Bethany Jackson, Vugar Ismaylov, Adeoye Oluwakanyinsola Debo-Aina, Findlay MacAskill, Francesca Kum, Meghana Kulkarni, Raveen Sandher, Anna Walsh, Ella Doerge, Katherine Guest, Yamini Kailash, Nick Simson, Cassandra McDonald, Elsie Mensah, Li June Tay, Ramandeep Chalokia, Sharon Clovis, Elizabeth Eversden, Jane Cossins, Jonah Rusere, Grace Zisengwe, Louisa Fleure, Leslie Cooper, Kathryn Chatterton, Amelia Barber, Catherine Roberts, Thomasia Azavedo, Jeffrey Ritualo, Harold Omana, Liza Mills, Lily Studd, Oussama El Hage, Rajesh Nair, Sachin Malde, Arun Sahai, Archana Fernando, Claire Taylor, Benjamin Challacombe, Ramesh Thurairaja, Rick Popert, Jonathon Olsburgh, Paul Cathcart, Christian Brown, Marios Hadjipavlou, Ella Di Benedetto, Matthew Bultitude, Jonathon Glass, Tet Yap, Rhana Zakri, Majed Shabbir, Susan Willis, Kay Thomas, Tim O’Brien, Muhammad Shamim Khan, Prokar Dasgupta

## Abstract

**Background:** Contracting COVID-19 peri-operatively has been associated with a mortality rate as high as 23%, making prevention vital.

**Objectives:** The primary objective is to determine safety of surgical admissions and procedures during the height of the COVID-19 pandemic using ‘hot’ and ‘cold’ sites. The secondary objective is to determine risk factors of contracting COVID-19.

**Design, Setting and Participants:** A retrospective cohort study of all consecutive patients admitted from 1^st^ March – 31^st^ May 2020 at a high-volume tertiary urology department in London, United Kingdom. Elective surgery was carried out at a ‘cold’ site requiring a negative COVID-19 swab 72 hours prior to admission and to self-isolate for 14 days pre-operatively, whilst all acute admissions were admitted to the ‘hot’ site.

**Outcome Measurements and Statistical Analysis:** Complications related to COVID-19 were presented as percentages. Risk factors for developing COVID-19 infection were determined using multivariate logistic regression analysis.

**Results and Limitations:** A total of 611 patients, 451 (73.8%) male and 160 (26.2%) female, with a median age of 57 (interquartile range 44-70) were admitted under the urology team; 101 (16.5%) on the ‘cold’ site and 510 (83.5%) on the ‘hot’ site. Procedures were performed in 495 patients of which 8 (1.6%) contracted COVID-19 post-operatively with 1 (0.2%) post-operative mortality due to COVID-19. Overall, COVID-19 was detected in 20 (3.3%) patients with 2 (0.3%) deaths. Length of stay was associated with contracting COVID-19 in our cohort (OR 1.25, 95% CI 1.13-1.39). Limitations include possible under reporting due to post-operative patients presenting elsewhere.

**Conclusions:** Continuation of surgical procedures using ‘hot’ and ‘cold’ sites throughout the COVID-19 pandemic was safe practice, although the risk of COVID-19 remained and is underlined by a post-operative mortality.

**Patient Summary:** Using ‘hot’ and ‘cold’ sites has allowed the safe continuation of urological practice throughout the height of the COVID-19 pandemic.

## Introduction

### Background

The spread of COVID-19, the respiratory disease caused by the virus Severe Acute Respiratory Syndrome Coronavirus 2 (SARS-CoV-2) was first reported in the United Kingdom in the week commencing 27^th^ January 2020 and was declared a pandemic by the World Health Organisation on 11^th^ March 2020 (1) (2). By 17^th^ March 2020 the British Government postponed all non-urgent surgical operations to reduce spread and reallocate resources to target the disease (3). Patients having surgical operations may be at increased risk of contracting COVID-19 due to the aerosol generated during anaesthesia and the procedure itself, which may mobilise pathogens as well as the immunosuppressive response to surgery (4). Post-operative pain limiting respiration and immobility can also make patients more susceptible to respiratory infections.

Pulmonary complications related to COVID-19 reported by the COVIDSurg collaborative, an international multicentre observational study found post-operative respiratory complications occurred in half of patients that had contracted COVID-19 peri-operatively (from 7 days pre-operatively to 30 days post-operatively) and found 30-day mortality to be 23.8%. (5) Given these outcomes it is imperative to prevent peri-operative contraction of the virus and our centre has developed ‘hot’ sites for acute admissions and ‘cold’ sites for elective admissions requiring patients to self-isolate for a minimum of 14 days and produce a negative COVID-19 polymerase chain reaction (PCR) swab prior to procedure.

The department described in this study is within a high-volume tertiary referral centre. Guy’s & St Thomas’ NHS Trust was at the centre of the pandemic, there were over 1400 cases of COVID-19 diagnosed and over 330 ventilated in intensive care throughout the pandemic, which is the most in the UK. Up to 2000 members of staff and 20% of the workforce took sick leave either unwell as a result of the virus or isolating. The current study helps to inform management in other centres with high incidence of COVID-19 in the community.

### Objectives

The primary objective was to determine the safety of the continuation of urological admissions and procedures through the height of the COVID-19 pandemic using ‘hot’ and ‘cold’ surgical sites. The primary outcomes were post-operative COVID-19 infection and mortality related to COVID-19. The secondary objective is to determine risk factors of contracting COVID-19 within our cohort to determine which patients are more at risk and assist further preventative strategies.

### Patients and Methods

#### Study design

An observational cohort study of all consecutive patients admitted at the height of the COVID-19 pandemic from 1^st^ March 2020 – 31^st^ May 2020 across both ‘hot’ acute sites and ‘cold’ elective sites was performed. Peri-operative infection, complications and mortality related to COVID-19 were reported to gauge the safety of the continuation of surgical procedures throughout the pandemic. Risk factors for developing COVID-19 infection within our cohort were determined using multivariate logistic regression analysis. Data is presented in line with the Strengthening the Reporting of Observational Studies (STROBE) statement for cohort studies. (6)

#### Setting

A single surgical department of a tertiary care referral centre in London, United Kingdom. Patients managed at the ‘cold’ site were ‘COVID-19 protected’, requiring patients to self-isolate for a minimum of 14 days and produce a negative Roche COVID-19 polymerase chain reaction (PCR) antigen swab within 72 hours prior to the procedure (7). ‘Hot’ site admissions were ‘risk managed’ and included all patients admitted acutely throughout the 3 months of the study, as well as all elective admissions prior to ‘cold’ site opening on 30^th^ March 2020. If patients were diagnosed with COVID-19 they were managed in isolation or in wards with only COVID-19 positive patients. All COVID-19 negative or not suspected patients were managed on wards without isolation but with personal protective equipment used. On both sites Public Health England personal protective equipment (PPE) guidance was followed; for all inpatient care fluid repellent surgical masks, apron, gloves and eye protection and for aerosol generating procedures filtering facepiece masks (FFP3), fluid repellent gown, gloves and eye protection were worn by healthcare professionals (8). Elective admissions for extracorporeal shock wave lithotripsy remained on the ‘hot’ site throughout with the same isolation precautions required prior to treatment.

#### Participants

All consecutive patients admitted under the care of the urology team. Patients were identified using a prospective consecutive dataset of admissions and surgical bookings.

#### Variables

Variables recorded were age, gender, ethnicity, COVID-19 positive (yes/no), complications related to COVID-19 (if positive), respiratory support required for COVID-19 (if positive), mortality (yes/no), hypertension (>140/90 mmHg), Charlson Comorbidity Index, operation performed, length of stay (in days).

#### Data sources

Data was retrospectively collected using a prospective consecutive dataset of admissions using trust-wide electronic patient records and morbidity and mortality data. Data was gathered electronically from surrounding hospitals when reported to have presented elsewhere.

#### Bias

There was a risk of underreporting due patients presenting in other trusts with COVID-19 infection or subclinical COVID-19 infection in the community.

#### Statistical Methods

Complications related to COVID-19 were presented as percentages of the overall cohort and number of patients that underwent surgical procedures. Risk factors for developing COVID-19 infection within our cohort were determined using multivariate logistic regression analysis. Gender, ethnicity hypertension, operation (yes/no) and COVID-19 status were analysed as binary variables, age (≤70 or >70) and Charlson Comorbidity Index as dichotomous variables and length of stay as a continuous variable.

#### Ethics

This service evaluation/audit was granted institutional approval by the Guy’s and St Thomas’ Hospital and the requirement for consent for data use that was anonymised before analysis was waived.

## Results

### Participants and descriptive data

A total of 611 patients, 451 (73.8%) male and 160 (26.2%) female with a median age of 57 (interquartile range 44-70) were admitted under the surgical team (table 1). Of these, 101 (16.5%) were admitted on the ‘cold’ site and 510 (83.5%) on the ‘hot’ site (table 1). Surgical procedures were performed in 495 (81%) (table 2).

**Table 1.**
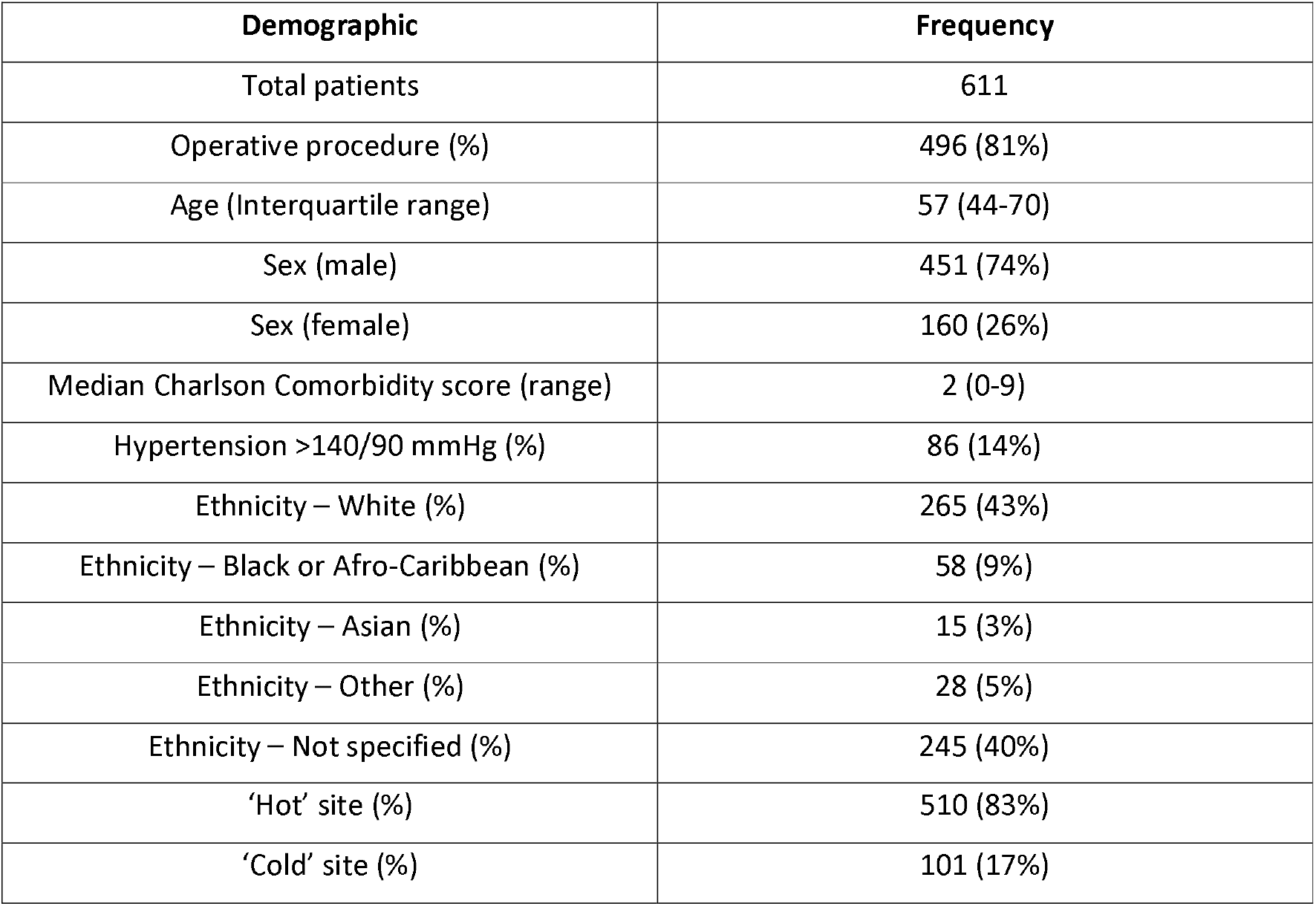
Patient Demographics.

### Primary Objective

Overall, COVID-19 was detected in 20 (3.3%) patients with 2 (0.3%) mortalities. Of the 495 patients that underwent surgical procedures, 8 (1.6%) contracted COVID-19 post-operatively with 1 (0.2%) post-operative mortality due to COVID-19. Of the 20 COVID-19 positive patients 17 were at the ‘hot’ site and 3 at the ‘cold’ site, 2 of which were detected pre-operatively and 1 post-operatively.

Supplemental oxygen and antibiotics were required in 4 patients that developed COVID-19 while another required additional methylprednisolone and awake proning but avoided intensive care (Table 3). There were 2 mortalities in patients that contracted COVID-19; one post-operative mortality following transurethral resection of bladder tumour at the ‘cold’ site with negative pre-operative COVID-19 swab that presented to a local hospital in respiratory failure 14 days following surgery and one patient with palliative metastatic bladder cancer and respiratory failure due to COVID-19 that did not have an operation.

**Table 3.**
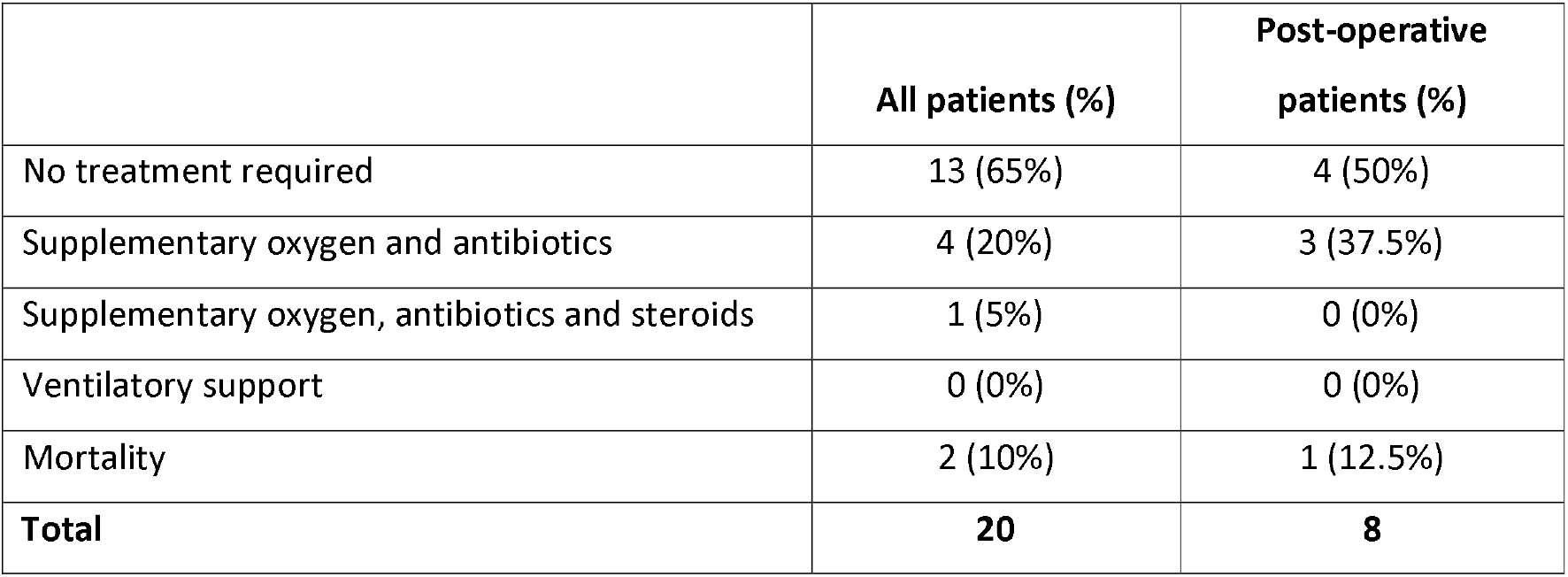
Management required for COVID positive patients.

**Table 4.** Risk factors of contracting COVID-19 following multivariate analysis. All odds ratios (OR) were adjusted for minimal variables.

| Variable | No. of COVID events | OR | 95% CI | P for Trend (for continuous variables only) |
| --- | --- | --- | --- | --- |
| **Age** |  |  |  |  |
| ≤70 | 8 | 1.00 | Ref. |  |
| >70 | 12 | 2.20 | (0.54-8.99) |  |
| **Gender** |  |  |  |  |
| Female | 4 | 1.00 | Ref. |  |
| Male | 16 | 1.18 | (0.38-3.65) |  |
| **Black or Asian Ethnicity** |  |  |  |  |
| No | 15 | 1.00 | Ref. |  |
| Yes | 5 | 1.83 | (0.57-5.86) |  |
| **Cancer** |  |  |  |  |
| No | 10 | 1.00 | Ref. |  |
| Yes | 10 | 1.13 | (0.44-2.91) |  |
| **Hypertension** |  |  |  |  |
| No | 13 | 1.00 | Ref. |  |
| Yes | 7 | 0.70 | (0.26-1.84) |  |
| **Charlson Comorbidity Index** |  |  |  |  |
| 0 or 1 | 1 | 1.00 | Ref. |  |
| 2 | 3 | 12.15 | (1.24-118.93) |  |
| 3+ | 16 | 15.24 | (2.00-115.77) |  |
| **Operation/procedure** |  |  |  |  |
| No | 4 | 1.00 | Ref. |  |
| Yes | 16 | 1.10 | (0.31-3.83) |  |
| **Length of stay (cont.)** | N/A | 1.25 | (1.13-1.39) | <0.000 |
| **Site of admission** |  |  |  |  |
| ‘hot’ site | 17 | 1.00 | Ref. |  |
| 'cold' site | 3 | 0.86 | (0.25-3.01) |  |

*OR – Odds ratio; 95% CI – 95% confidence interval*

### Secondary Objectives

On multivariate analysis, length of stay was the associated with contracting COVID-19 in our cohort (OR 1.25, 95% CI 1.13-1.39). Patients with higher Charlson Co-morbidity Index of 3 or above also looked to be more at risk (OR 15.24, 95% CI 2.00-115.77) pointing to comorbidity as a risk factor. Patients having surgical procedures were not at higher risk of contracting COVID-19 compared to those that did not (OR 1.1, 95% CI 0.31-3.83).

## Discussion

In our cohort the risk of contracting COVID-19 remained low (3.3%) throughout the peak of the COVID-19 pandemic in the United Kingdom over both ‘hot’ and ‘cold’ sites. Given that the hospital was at centre of the pandemic in London in a severely affected area, this model was a relatively safe method of continuing to deliver surgical care. One challenge going forward will be dealing with increased waiting lists and delivering safe care on a larger scale. There was a significant reduction in the elective service during the time of the study; the described department typically has between 600-1000 inpatient episodes monthly, in contrast to the 200 per month described in this series. This is a worldwide phenomenon with up to 2.8 million surgeries worldwide expected to be delayed cancelled as a result of COVID-19, making it vital to find a safe method of carrying out operations following the pandemic. (9) The ‘cold’ site model has been safely described elsewhere, and reports from another ‘cold’ site in London have reported a 2% rate of post-operative COVID-19 infection, with only 0.2% of their cohort reporting Clavien-Dindo grade 3 complications related to COVID-19. (10) The current study supports the practice of ‘cold’ sites in areas still suffering from high rates of COVID-19.

Post-operative mortality related to COVID in our cohort was 12.5% (1/8) which is lower than the figure of 20.5% reported in China and 23.8% described by the worldwide COVIDSurg collaborative (11) (5). Interestingly, apart from the reported mortality there were no Clavien-Dindo 3 or above complications in the current series related to COVID-19 although 37.5% (3/8) of post-operative patients required supplementary oxygen and antibiotics. Over the whole cohort there were no patients from our department that required ventilatory or intensive care support apart from the two mortalities. The two patients had significant comorbidities, with Charlson Comorbidity Index scores of 7 and 8 respectively. Multivariate analysis demonstrated that those with greater comorbidity were at a higher risk of contracting COVID-19, so surgery in these patients must be taken with greater planning and caution in areas with high COVID-19 prevalence.

An interesting finding of the current study was the association of length of stay with increased COVID-19 infection. It is unclear whether the patients requiring longer admissions are more at risk, pointing to hospital-related transmission or if COVID-19 infection led to a prolonged admission. However, as no patients required intensive care and only 5 required supplementary oxygen and antibiotics, it is unlikely that the majority of patients with COVID-19 had an extended stay due to the virus. The three patients with the longest stay with COVID-19 did not require oxygen and were likely to have been infected while in hospital. This suggests that early discharge where clinically appropriate may aid the reduction in hospital-related COVID-19 transmission. In practice surgical departments have attempted to do this and admissions have been reported to have a reduced length of stay during lockdown when compared to previously (12). Risk factors for a more severe course of COVID-19 have been described and include age 65 and older, living in a nursing home or long term care facility as well as chronic lung disease (13). The rationale for surgery in these patients is particularly important, in the described department all ‘cold’ site surgeries were discussed at a multidisciplinary team meeting and prioritised according to The Royal of College of Surgeons guidance and added to waiting list or deferred according to clinical need. (14) Surgery in such patients must be taken with extra precautions and patients and the family informed of higher risk of mortality.

Benefits of ‘cold’ and ‘hot’ site work included the reduced cross-contamination of COVID-19 infection from the acute site. Global guidance for surgical care during the COVID-19 pandemic has suggested patients should be cared for by COVID-19-specific surgical teams if possible, rather than those who are also seeing uninfected patients which was followed in our site. (15) To reduce cross-contamination, members of the surgical team were not permitted to visit the ‘cold’ site on the same day as the ‘hot’ site although this did not come into effect until a few weeks into the opening of the ‘cold’ site as the ‘cold’ site evolved. Across both sites appropriate PPE guidance was followed in inpatient and outpatient areas according to Public Health England Guidance. (8) This has been a successful model in our centre where two adjacent separate sites were available, however not all centres have this facility and separation may have to occur within a single site.

One limitation of this study the possibility of under-reporting of post-operative COVID-19 infection as a result of patients presenting at other hospitals or sub-clinical infection in the community without presenting to hospital. Asymptomatic shedding of the virus can occur and may account for up to 60% of cases. (13) To limit this, the authors contacted surrounding hospitals where patients or doctors reported presentations in other sites, as is the case with the post-operative mortality reported. The observational nature of this study meant that the authors did not request patients to have routine post-operative COVID-19 PCR swabs which may have increased the number of post-operative infections detected. On the other hand, post-operative COVID-19 infection is not necessarily causative from the admission as COVID-19 could have been obtained in the community in patients discharged from care. In addition, the ‘cold’ site was not set up at the beginning of the time described and was a response to the pandemic, coming into effect on the 30^th^ March 2020. However, the authors decided to include all patients in March 2020 as this gives a more rounded picture of the height of the pandemic. Not all patients at the ‘hot’ site were tested for COVID-19 on admission although all ‘cold’ site patients were. Routine testing on the ‘hot’ site changed through the time of the study, initially only symptomatic patients were being tested, while later all emergency admissions were tested. Due to the collection of information using electronic records, ethnicity was not recorded in all patients with 40% not specified on records, meaning conclusions about ethnicity based on this dataset were limited. This is an important cohort as black and Asian patients have been associated with worse outcomes when compared to white patients (16) (17). Due to the low number of mortalities and serious complications related to COVID-19 we have been unable to make conclusions of whether risk factors are predictors of morbidity or mortality. Whilst this is a single centre study, our unit is a high volume tertiary urology and oncology centre and outcomes here may be able to guide other centres considering ‘cold’ and ‘hot’ site models.

## Conclusions

Continuation of surgical procedures using ‘hot’ and ‘cold’ sites throughout the COVID-19 pandemic was reasonably safe and enabled patients undergo important procedures, although the risk of COVID-19 remained and is underlined by a post-operative mortality. We identified risk factors that could limit transmission, notably reducing length of stay.

## Data Availability

This service evaluation/audit was granted institutional approval by the Guys and St Thomas Hospital and the requirement for consent for data use that was anonymised before analysis was waived.

## Appendix 1

**<u>Table 2. All procedures completed (see separate attachment)</u>**

